# Awareness of Intimate Partner Violence among Faith-Based Leaders of Migrant and Refugee Background in New South Wales, Australia

**DOI:** 10.1101/2024.07.17.24309491

**Authors:** Naomi K. Kayesa, Olayide Ogunsiji

## Abstract

**Purpose:** Intimate Partner Violence (IPV) is a family violence issue requiring public health system interventions. Migrants and refugees experiencing IPV often rely on their Faith-Based Leaders (FBLs). However, the role of FBLs is under-reported. This study aims to report findings on awareness to IPV among FBLs of migrant and refugee backgrounds.

**Methods:** This qualitative study engaged 12 FBLs in face-to-face interviews. Thematic analysis revealed three themes namely: Awareness and understanding of IPV; Being the first point of call; Awareness of available services for IPV.

**Results:** All FBLs described IPV as relational and identified the different IPV types. Sexual violence did not resonate as IPV by eleven participants. Explanations for being the first point of contact centered around victims’ quest for a win-win outcome and in-accessibility of formal services.

**Conclusions:** IPV specific training, collaboration between FBLs and formal services working with victims of IPV is essential in supporting IPV victims.

## Introduction

Intimate Partner Violence (IPV) is a form of domestic violence (DV) issue with significant physical, emotional, sexual, and psychological health consequences. It is on the increase and affecting most families, including migrant and refugee families [1, 2], hence a public health issue. Of the seven million migrants making Australia their home, 252,000 (30%) live in New South Wales (NSW) [3] and represent an integral part of the population experiencing various forms of domestic violence. Although it is still unclear (in the Australian context) whether migrants encounter domestic violence more than other populations, it is known that they are significantly more vulnerable due to their migration status [4, 5]. Factors directly associated with IPV among migrants and refugees include pre-arrival traumatic experiences, acculturation, change in gender roles, immigration status and social isolation among others [6,7,5].

Faith based leaders play an integral role in addressing IPV among migrants and refugees [8,9]. When faced with IPV, migrants and refugees gravitate towards communities with whom they share a common background and religious beliefs, in particular FBLs. Despite these leaders being the preferred source of support, their voices are yet to be heard about their awareness and involvement in issues relating to IPV. Little is known about their accessibility to NSW formal services in equipping themselves to effectively work with the migrants and refugees encountering IPV.

This article reports on awareness of IPV among FBLs of migrant and refugee background in NSW. It is important to gain this insight to identify knowledge gaps and develop relevant strategies for addressing these gaps.

## Background

Buchanan (2011) [10], described FBLs as front-runners in faith-based organizations. Also known as religious leaders, they are at a vantage position in raising IPV awareness and influencing attitudes, behaviors, social values, and practices through faith-based teachings. Their members approach them for counsel about IPV related issues and various aspects of their lives. For decades, FBLs have assumed a range of roles including international conflict resolution [11], transformative family processes [12] and have proved to facilitate refugees’ resettlement.

It is documented that communities with a history of conflict and a myriad of traumatic experiences face an extension of violence in their homes [13–16]. Sociological theories also assert that violence in families is highly associated with de-masculinization among most post-conflict males, rendering women and children the commonest victims of domestic violence [17, 18]. Besides conflict, migrants, and refugees in general face a multitude of challenges that range from navigating new systems, learning a new language, culture shock and securing new jobs [19]. While it might be relatively easier for local citizens of the country to navigate their way around IPV, the same cannot be said for migrants and refugees. They often lack information regarding existing resources, policies, and services to protect themselves from effects of IPV [20]. Besides, empirical evidence reveals that migrant groups suffer discrimination at the hands of their communities due to cultural beliefs that emphasize the sacredness of marriage over domestic abuse [21]. Further to this, law enforcers focus more on visa status, child welfare and other migration issues rather than offering support against IPV [13, 22]. All these explain why victims seek counsel from informal sources such as FBLs.

It is apparent that FBL of migrant and refugee backgrounds are usually the first contact for most migrants seeking help with regards to IPV. In their study, Bermúdez, Kirkpatrick, Hecker, and Torres-Robles (2010) [23] confirmed that FBLs are typically the first point of contact for migrants when faced with IPV. According to Bermudez et al. (2010) [23], only 6% of the participants in the study indicated reporting to other professionals while 21% reached out to a religious leader first. The participants explained that faith-based institutions and leaders provided better informal services, free of fear and intimidation even for undocumented migrants. Additionally, they found that victims of IPV were most likely to reach out to an FBL as it was easier to talk to someone who spoke their language and understood their cultural values. Along the same vein, Choi and Cramer (2016) [24] found that 92.7% of immigrant clergy counselled individuals and families facing IPV, demonstrating the multitude of IPV victims that are constantly in contact with FBLs.

However, the situation with the FBLs in NSW and what they offer in supporting their members experiencing IPV is not clear in the literature. This is important to know to identify gaps in areas of support for their members.

Meanwhile, there are instances where Christian FBLs have been the last resort for migrants and refugees living with IPV [19,25]. In a study conducted in the United States of America (USA), victims preferred reaching out to their elders first [19]. It was only when the results of the support offered by elders were unsatisfactory or when the abuser re-offended, would the victims seek advice from religious leaders. In that same study [19], some victims consulted their relatives first to keep the violence within the family or when faith-leaders could not be reached. Similarly, Islamic FBLs in the USA self-reported that they were not the first point of contact [25]. Due to previous negative experiences whereby, FBLs prioritized traditional and cultural beliefs over the protection of victims, they ceased to be the first point of contact for IPV victims [25]. Given this inconclusive situation, it is important to gain insight from FBLs in NSW, Australia (where a predominant population of migrant and refugee families live) with regards to where they are positioned when their members experience IPV.

Awareness of IPV and capacity to address it by some FBLs of migrant and refugee backgrounds have been questioned by some members of their organizations who contacted them for support [19]. While most men have expressed satisfaction with the way IPV issues were handled by FBLs, women felt blamed and judged by the same [19]. Possible explanations ranged from a lack of understanding of IPV, and the lack of appropriate skill set to provide efficient support to those victimized. Contributing to this is the migrant status of the FBLs themselves. Findings from a literature review revealed that only 16% (n=24) of migrant FBLs were adequately equipped and knowledgeable to deal with IPV when approached by victims [26]. This is due to several reasons which range from the FBLs lack of awareness of the systemic procedures surrounding IPV in their new countries, to faith teachings that over-emphasize the need for women to be submissive to their husbands. In the process of reinforcing migrant beliefs and cultures alongside biblical references, the message is misconstrued, and judgement is biased in favor of the abuser [27, 28]. Empirical studies have yet to clarify the situation for NSW. Through this empirical study, we give FBLs of migrant and refugee background in Australia a voice on their awareness of IPV and their perceived capacity to support members that approach them.

## Materials and Methods

### Purpose

The aim of this paper is to report awareness of IPV among FBLs of refugee and migrant background in New South Wales, Australia.

### Design

This qualitative study utilized narrative inquiry approach to gain insight into FBLs’ awareness of IPV. According to Clandinin and Connelly (2000) [29] “narrative inquiry is stories lived and told” (p.20). Faith Based Leaders are the first point of contact for migrants and refugees living with IPV. They are in the best position to talk about what they know and their level of awareness of IPV. Gaining this insight will assist in identifying knowledge gaps and inform targeted material for raising awareness of IPV among these leaders of migrant and refugee background.

### Participants

Participants for this qualitative paper were drawn from those who responded to an online questionnaire in a bigger study titled “awareness and response of IPV among FBLs of migrant and refugee background”. At the completion of that questionnaire, participants who were willing to provide deeper insight and rich text data were invited to provide their names and contact phone number or email address. For de-identification purposes, pseudonyms have been used in this paper.

### Inclusion criteria

To participate in this study, the person had to self-identify as a faith-based leader, 18 years and over, of migrant or refugee background, with experience of dealing with IPV among the people he/she is leading and be willing to participate in the study.

### Sample size

A total of 12 FBLs who indicated their willingness to participate in face-to-face or telephone digitally recorded interviews (by providing their names and contact phone numbers or email address) took part in this qualitative study. However, pseudonyms have been used in this paper (intext and information table in appendix).

### Data collection

Consistent with narrative inquiry, data collection for this study was through face-to-face or telephone digitally recorded interviews. Prior to the commencement of interviews, participants were provided with a participant information sheet which contained detailed information about the study. They were encouraged to ask questions and clarify any content of the participant information sheet. After the participants verbalized their understanding of the focus and what is expected of them in the research, they were offered consent forms for completion. Meanwhile, those that participated in telephone interviews provided verbal consent. Furthermore, participants were reminded that their participation in the study was voluntary and that they could decide to terminate their participation in the study at any time or decline to respond to any question if they wish to, without providing explanation. Face-to-face interviews went from 45-60 minutes at mutually suitable venues nominated by researchers and participants. All interviews took place at a time convenient for researchers and participants.

A total of seven interviews were conducted by telephone, while five were face-to face. Two researchers were responsible for data collection and each of them conducted an average of four interviews. In narrative inquiries, researchers “situated individual stories within participants’ personal experiences, their jobs, their culture and context in general” [30] Questions about the participants’ gender, religion and self-introduction were asked and findings are presented in the demographic Table 1. The interviews started with an open-ended question, “Can you please tell me everything you know about IPV (domestic violence)?” Depending on their initial response to the question, further probing questions included “You have presented your understanding of IPV, can you please tell me the different types that you know? “Can you please tell me the different types of IPV/DV that you have handled among the people you are leading?”

**Table 1.** Demographic Characteristics of Faith Based Leaders.

Demographic Characteristics of Faith Based Leaders
| Name | Gender | First reporter | Self-introduction | Religion |
| --- | --- | --- | --- | --- |
| JJ | Male | Men and women | Pastor, migrant, community leader | Christianity |
| SS | Female | Women | Women's leader, Migrant | Christianity |
| AA | Male | Men and women | Community leader, Marriage celebrant, Justice of Peace Senior Pastor, Refuge | Christianity |
| MM | Male | No direct experience of dealing with IPV | Faith based leader | Christianity |
| AA | Female | Men and women | Senior Pastor | Christianity |
| FF | Male | Women | Pastor | Christianity |
| PP | Male | Most of the time women | Pastor | Christianity |
| MO | Male | Men and women | Pastor | Christianity |
| SA | Female | Mostly women | Religious leader, background; parenting and family coach | Islam |
| OO | Male | Men and women. | Religious leader, used as an interpreter | Islam |
| JJ | Male | Male and Female 50-50 | Community leader | Islam |
| II | Male | Mostly female | Religious leader | Christianity |

The interaction between the researcher and the participant during data collection was such that the researchers played the roles of listener and questioner. In this case, the researchers asked the questions from their respective participants and then listened while the participants presented their responses. After this, follow-up questions were then asked, and this process continued until the end of the interviews. The interview data was transcribed verbatim.

### Data analysis

Informed by Van Manen (1990) [31], directions with regards to qualitative data analysis, both researchers read the interview transcripts to immerse ourselves in the data. Once we got the sense that is being made in the data, we looked for reoccurring sentences, concepts, and phrases. These were mapped out to refine categories and to assign meaning to the data. The refined categories were further developed into emerging themes. Three themes emerged-*awareness and understanding of IPV; Being the first point of call; awareness of available services for IPV*.

### Ethics approval

Ethics approval was obtained from an Institutional Human Ethics Committee.

## Results

A total of 12 FBL from Christian and Islamic religions took part in this study. Seventy-five percent (n=9) of these participants were male while 25% (n= 3) were female. The participants were migrants and refugees of African and Middle-Eastern backgrounds. Their years of faith-based leadership ranged between 4 and 20 years in Australia and overseas. In addition to being FBLs, five of the participants included some other appellations in their self-introduction such as community leaders, marriage celebrants, Justice of peace, parenting and family coaching. Eight participants migrated to Australia while the remaining four arrived in Australia as refugees. A summary of the participants’ characteristics is provided in Table 1(see Appendices). Pseudonyms have been used for de-identification purposes.

### Awareness and understanding of IPV

All the participants had a good understanding of IPV. They were aware that IPV is usually experienced by someone in a very close and mutual intimate relationship “…so there is a relationship, there’s a mutual relationship along the lines” (AA). This relationship, according to most of the participants occurs between married male and female “…between a married couple. A husband and awife” (MM) and for some (n=2) it could be between “partners, you know, fiancé…” (AA) or simply “household members” (FF). They spoke about the settings within which IPV occur, that it occurs within the family setting “…occurs within the family setting. It could be at home, or it could be somewhere where the family’s, having all the families together” (AA).

The participants variously described IPV as abuse, neglect, harassment, and deprivation of rights and needs. Majority (n=8) described it as an abuse whereby one partner makes the other partner feel valueless “…. it is when one partner abuses another partner making them feel as if they are of no value” (SS). It is “…any incident or actions by either partner that causes some disturbance within the relationship itself” (PP). They noted the tendency to assume that IPV is only physical in nature “…when you talk of violence, you’re not only talking of physical violence from my understanding but even verbal abuse and even harassment” (AA). They emphasized that this abuse could manifest in various forms “this abuse can take different form, such as physical, verbal, emotional, economical, sexual abuse as well as neglect” (AA). “I want to say of violence it could also be in the area of neglect” (SS). The issue of deprivation of rights and needs came out clearly in the explanation of two of the participants. SS, perhaps due to her professional and specific training in parenting and family issues, provided a more sophisticated explanation of IPV. According to her, IPV is “manipulation or denial or dismissal or abuse of the right and need to the partner” (SA) while Omar emphasized that when one is “transgressing by words or by actions or by not fulfilling the rights from one party to the other”, this is IPV.

According to the participants, IPV is behavioral “…intimate partner violence, it’s a behavior with an intention to negatively impact and hurt another partner” (JJ), “…it’s doing things that would hurt the other partner” (SS). That it always starts from somewhere and progressively worsens with time “it’s something that is consequential. It’s something that has a beginning and then it escalates” (OM).

### Being the first point of call

All the participants acknowledged themselves as the first point of call for migrants and refugees living with IPV; “most cases coming to us as faith-based leader is their first point of call” (JJ), “when this violence occur, the first point of contact is us (AA). Prominence is the reason for the FBLs’ being the first point of call for the migrant and refugees bringing about a win-win outcome from the IPV experience. According to the participants, because they are of refugee and migrant background like the people they are leading, there is the general belief that they are the people who preach peace, reconciliation and are good examples within their families. Faith-Based-Leaders of refugee and migrant background understand their cultural context, the importance of family, many times speak the same language and often having resided in Australia longer than the victims of IPV:

“The reason why they come to us first is that they want a win-win outcome. They are not stupid, they know they could go to the police, but African women always consider the impact of reporting their husbands to the police on their family and their children, this is more important to them. They don’t want to lose; they don’t want to lose their marriage. So, because of that they would prefer to firstly come to the FBLs and when you are able to resolve it, a lot of them have been resolved like that and they are doing fine now. When you are able to do that, yes everybody’s happy. It’s the win-win situation” (JJ).

“And I mentioned to the community that I’m a Sheikh of reconciling and not a Sheikh that issues divorce statements for people easily. We give them a chance; we give spouse a chance to reconcile before they have their divorce in place and so many cases, they just need this advice they just need to delay and think about what they are doing and, in most cases, they refrain from divorce because they had this advice of not going ahead with the divorce just they have to think more about” (Sheikh).

“Because she feels more comfortable speaking to someone from the same background, same language, therefore understanding where they come from. When they raise the point, some cultural point that people could understand much effectively than someone who is strange to that culture” (MO).

Despite that FBLs’ capacity to address IPV appropriately involves a different skill set wider than simply understanding IPV, most of them reported to have taken up the role of IPV resources persons by virtual of being leaders in their respective religious institutions. Being religious leaders, the people that they are leading are also religious and prefer biblical and qur’anic approaches to resolution of their family issues. Many often referred to as spiritual leaders, they are trusted and respected members of the community and the go to person in difficult times, “…when they have this problem (IPV) they are looking for trust and trust-worthy person they can go to. And I’m representing the Muslim value and I’m representing Islam to them” (Sheikh). Some of these FBLs are seen as advocates for people living with IPV in their communities:

“Why do they come to me? yeah because I advocate for them, I publicly talk out, to educate women about it. I talk out against it (IPV), I talk about that we need to be encouraging our Sheikhs and Mashaykh, which is basically our FBLs to have a better understanding of it because they are usually the first source where a woman will go to seek a Muslim or will go to just seek advice regarding her marriage” (SS).

### Awareness of available services for IPV

In response to the question of their awareness of available services for IPV, the participants stated that there are a lot of support services “outside there” (JJ). All the FBLs in this study were aware of available formal support services. They readily mentioned police; “I think the first line of support from the moment that the violence is involved, I think the first fore line is the police and refuge centers. I also know there’s a lot of NGOs out there especially you know for women, a women’s shelter. I think there are a few of them” (FF).

“They actually have men’s shelter out these days” (PP), “…Family Assistant Office, Websites, I mean we’ve got websites, there’s many websites on, I can’t think off hand, I’ve got a list of them and that…. but there are different websites on domestic violence and legal organization” (SA). The majority explained that they know where to find necessary information if required. Meanwhile from the participants’ stories, it was gathered that there is a difference between awareness of available services and making referral to those services. They identified several reasons for their referral or otherwise to these services. All the participants identified the value of support services provided by the police and women’s refuge particularly in protecting the victims and preventing untimely death “I don’t have anything against…. other support organizations, they may be necessary to protect the victim and to prevent violence that can be deadly” (JJ). They expressed their willingness to seek support from available formal services for cases they refer to as critical; “…we might have to refer to the police if it is something that is critical to different organizations if we have to” (AA). “We have of course if it’s really life threatening or physical threatening or even more just people at risk, there is referral of the police” (MM).

Majority of the participants who had never referred individuals and families to the formal support services predominantly spoke about their capacity to resolve a few IPV incidents that were reported to them; “they (members of the congregation) know that when the pastor is involved, it’s going to be resolved prayerfully and amicably, which has been happening” (AA). For many non-referral and unwillingness to refer, very importantly is the shared perception of the police by FBLs and their followers. The participants explained that there is the general perception that the police and other available services such as the refuge centers do more damage than good to families:

“The reason is most woman believe that the outcome and I think so, I tend to agree with them they will tell you that the only reason why I have not taken my wife or my husband to the police is that the outcome is not always good. That they break into the home. They separate them. In the process to protect the victim they sever the relationship and they said they believe that they destroy their marriages… (JJ).

As a result, they do not want to be accused of suggesting avenues which the contending families do not trust for a positive outcome. The participants explained that they prefer that the victims take their personal initiative to contact the formal services rather than being the conduit connecting them. The participants expressed their commitment to ensuring that they mediate in families living with IPV. However, if this mediation is not adequate to resolve the issue, it is then left for the partners to decide about seeking formal support: “let them do it their way, not that ‘oh, pastor sent me to take you there’. “It’s the pastor that said that you go, take him to the police. They will take themselves to the police eventually, but it shouldn’t come from me as a faith-based leader, as a pastor” (JJ).

Some of the participants considered themselves lucky in that they had not had any instance where they needed to refer any of their members to the external organization or support. Rather, they have had members who had referred themselves to these external support systems “…what the members have done is to refer themselves directly to external organization. For instance, we have a couple who reported themselves to the police” (AA). In instances like this, AA reported a negative outcome: “I remember one of the members saying she (the member’s wife) reported me to the police, she spoilt my record and that is why I haven’t got any job until now… See that is the point we don’t want it to get to if she hasn’t reported to the police” (AA).

Furthermore, these FBLs were aware of personal circumstances, such as visa status that surround the lives of some of their members: “Sometimes we send them to Centrelink if they have permanent residency or if they have a citizenship. But if they have a temporary visa or are refugees, or on a student visa and he kicked her out of the house, where does she go?” (Sheikh).

Therefore, they would not make referrals to formal support services. One of the participants cited a case of her member who reported himself to the police after beating his wife and afterwards called his pastor. The immediate reaction from the FBL was “why did you do that? You haven’t got your papers. Why did you go ahead and do that? You didn’t tell me you are going to call the police” (AA).

## Discussion

Our findings from this study revealed that FBLs of African and Lebanese background had a good understanding of IPV. This is similar to the Korean American Church Leaders who also expressed their understanding of IPV [24]. Our participants spoke about the relational nature of violence and the settings within which it takes place. This is consistent with the Korean American Church leaders, who reported that IPV occurs among people in marriage relationship. However, in their understanding of the types of IPV, all the participants except one in our study, did not identify sexual abuse as IPV. This is different from previous studies among Christian and Muslim faith communities [19, 24]. The non-inclusion of sexual abuse in most of the definition of IPV in our current study may be due to cultural reasons rather than religious. Faith-Based Leaders need to recognize sexual abuse as IPV to include it in their conversations with couples living with IPV. Targeted information about awareness of IPV among FBLs of migrant and refugee background needs to put sexual abuse in the forefront of discussions.

The FBLs of African and Lebanese backgrounds in this study self-acknowledged the integral role they play in addressing IPV among migrants and refugee communities. This is similar to the important roles previously reported for FBLs in influencing and changing peoples’ behaviors [10], their efforts in international conflict resolution [11], and their unique contribution to refugees’ local integration and meeting their socio-economic needs [32]. The participants in our study added to this list by mentioning instances where their interventions have yielded positive outcomes for couples living with IPV. They emphasized that they are the most visited and the first point of contact similar to the Latinos in the USA [23]. Meanwhile, this study did not compare the rate at which victims contacted female versus male FBLs given the unequal representation in participants, nor did it explore the gendered interventions to articulate how this translates to satisfying experiences for women living with IPV. This area requires further exploration for migrants and refugees in Australia to establish if disclosures to female FBLs is similar or different from disclosures to male FBLs, how male FBLs address violence perpetrated by the women’s husbands, and whether interventions are directed to male perpetrators of violence, or to couples as a unit experiencing IPV. This is particularly so, given that West African migrant women in the USA reported that FBLs often maintain a strict gender hierarchy, leaving many of them feeling unheard and dissatisfied [19].

The study results also highlight the ambiguity surrounding the official training and required capacity of FBLs in dealing with IPV. There is no evidence that the FBLs can accurately assess other risks or are aware of the negative impacts of IPV on children. We therefore suggest that capacity development training for FBLs focuses on the dangers of strict gender hierarchical approaches with emphasis on meaningful and satisfying interventions among other essential risk assessments.

A degree of mistrust of the police and official services was also noted in the FBL as evidenced by their reluctance in reporting IPV incidents to authorities especially when IPV perpetrators are not documented citizens of Australia. This speaks to the need for change in police interventions especially when the risk of lethality is high. There is need for culturally integrative approaches by police when responding to IPV amongst migrants and in processes that aim at improving FBLs’ capacity to effectively respond to IPV. Ideally, capacity building should be bi-directional as racial profiling by police and other police brutality myths among migrant FBLs could interfere with capacity building and relationships between the two parties. In view of this, the Government and agencies that have the responsibility for prevention and eradication of IPV among migrant and refugee population need to recognize these challenges and deliberately build these bridges to facilitate the capacity of these FBLs to support their members.

Important reasons for the FBLs being the most preferred and first point of contact for women living with IPV were enumerated by the participants in this study. The quest for a ‘win-win’ outcome and the FBLs’ understanding of the migrant and refugee women’s context were prominent among these reasons. This understanding of the migrant and refugee-specific context was also reported in a literature review of IPV among migrant population in the USA [24]. Being migrants and refugees, and most times of similar cultural background to their members, FBLs are considered to have a detailed understanding of what it means to be a migrant or refugee. These FBLs have insights into the cultural explanation that surrounds IPV and the fear of deportation due to visa status. This affirms FBLs as natural resources to support victims of IPV in migrant communities. This situation then presents the urgency for capacity building for FBLs to effectively support this vulnerable population.

Interventions that focus on fostering healthy relationships are more likely to be well-received among the migrants and refugees. The approach of the interventions must be Bible or Quran centered.

The FBLs’ stories suggest that people living with IPV have a specific outcome in mind, which is to retain their family. This is different from previous studies which indicate that “the very assumption that FBLs will ask them to keep families together rather than split them” is one of the very reasons some women delay contacting FBLs [33]. It is possible that the IPV experience for many victims in migrant and refugee population indicates that they would rather separate from abusive husbands. There are inconsistent findings from research on how much consideration is given to the stability of the family in the face of severe IPV, more research is proffered in the future.

The participants in this study expressed their awareness of available formal services for women living with IPV. Among the services they mentioned were the police, refuge centers and family assistance office. This is similar to the services mentioned by [5]. However, the participants’ stories revealed that there is a difference between availability and accessibility. The participants stated that they considered the formal services to be meant for protection of victims and prevention of violence-related untimely death. They emphasized that they put the severity of the IPV cases into consideration and that only critical cases are referred to the police. This is because of the negative perception of the police held by many FBLs and their followers. Some of these FBL even expressed instances of their followers’ having negative experiences with the police. We suggest avenues or interventions to clear the myths about the image of the police among migrant and refugee populations.

It is evident from our study that the FBLs of migrant and refugee background are willing to continue to support their followers living with IPV. Attention is thereby required to develop and build their capacity to do so. It is important to invest in services that people will use, thereby achieving a win-win outcome. Additionally, the adoption of culturally integrative models that have proven to effectively reduce IPV in migrant populations can be incorporated to harness the relationships between informal and formal services as they collectively facilitate FBL’s capacity in addressing IPV in minority populations [34]. Based on the findings of our study, there is an overwhelming potential for FBLs to prevent IPV among their members and support those living with it.

## Conclusions

There is significant evidence that FBLs play an integral role in shaping behaviors and ways of thinking among populations they lead. What is clear from the findings of this study is the lack of IPV services and training available which are faith-based and appropriate. It is important to note that interpretations of religion and socio-cultural factors play a crucial role as moderating factors of abuse. Negative perceptions of police involvement by victims of IPV heighten the role of the FBLs in resolving such matters, however, they are ill-equipped and under resourced. Lack of resources and intervention programs have not allowed FBLs to maximize their potential in the migrant or refugee communities facing IPV. Mainstream services are not inclusive and do not meet the needs of IPV victims of migrant and refugee background as they are not culturally sensitive and lack religious consideration. More critical is acknowledging the complexity of IPV particularly in migrant and refugee communities. Not only that, but also working towards building capacity for both FBLs and formal services to better deal with this complexity.

It is worth investing in resources and funding for intervention programs for FBLs of migrant and refugee background for them to fully support and educate their followers. In this instance developing a primary prevention program will serve to reduce numbers of IPV cases by dispelling myths leading to the exponential growth of IPV. The findings of this study revealed the importance of FBLs to their followers and the gap in culturally appropriate services and counselling in addressing IPV.

## Data Availability

All data is contained in the manuscript

